# Effectiveness of quarantine measure on transmission dynamics of COVID-19 in Hong Kong

**DOI:** 10.1101/2020.04.09.20059006

**Authors:** Hsiang-Yu Yuan, Axiu Mao, Guiyuan Han, Hsiangkuo Yuan, Dirk Pfeiffer

## Abstract

The rapid expansion of COVID-19 has caused a global pandemic. Although quarantine measures have been used widely, the critical steps among them to suppress the outbreak without a huge social-economic loss remain unknown. Hong Kong, unlike other regions in the world, had a massive number of travellers from Mainland China during the early expansion period, and yet the spread of virus has been relatively limited. Understanding the effect of control measures to reduce the transmission in Hong Kong can improve the control of the virus spreading.

We have developed a susceptible-exposed-infectious-quarantined-recovered (SEIQR) meta-population model that can stratify the infections into imported and subsequent local infections, and therefore to obtain the control effects on transmissibility in a region with many imported cases. We fitted the model to both imported and local confirmed cases with symptom onset from 18 January to 29 February 2020 in Hong Kong with daily transportation data and the transmission dynamics from Wuhan and Mainland China.

The model estimated that the reproductive number was dropped from 2.32 to 0.76 (95% CI, 0.66 to 0.86) after an infected case was estimated to be quarantined half day before the symptom onset, corresponding to the incubation time of 5.43 days (95% CI, 1.30-9.47). If the quarantine happened about one day after the onset, community spread would be likely to occur, indicated by the reproductive number larger than one. The results suggest that the early quarantine for a suspected case before the symptom onset is a key factor to suppress COVID-19.

## Introduction

The coronavirus SARS-CoV-2 outbreak, originally occurred from Wuhan, China, in December 2019 has caused a global pandemic in March 2020 [1, 2]. Regions in East Asia, such as Hong Kong, Taiwan, Korea and Japan, all faced an extremely high risk of community outbreak due to a massive number of travellers from Mainland China during the Chinese new year [3, 4]. However, comparing to many other countries until now (March 2020), the coronavirus outbreak was still considered limited in Hong Kong and Taiwan [5]. The low number of transmission can be due to a successful public health control strategy. To understand how coronavirus can be contained in these places is of great importance to limit the global spread of the virus happening right now.

Presently, nonpharmaceutical interventions including both mitigation and suppression strategies have been proposed to control the outbreak [6]. Suppression strategies were often intensive that can be challenging to fully implemented in many countries such as transportation restriction or city lockdown [4, 7]. How Hong Kong successfully prevent the outbreak without shutting down most of the public services thus offers us a different perspective on how to choose a containing strategy.

As most countries, quarantine and border control policies were taken in order to stop the spread of the coronavirus from Wuhan. During the Chinese New Year festival, which started on 24 January in 2020, more than one million of travellers (some were Hong Kong residents) arrived Hong Kong from Mainland in a week [8]. Local spread occurred starting from this critical moment when only few initial travellers who carried the disease. Therefore, infected individuals were reported almost every day with travel history from Wuhan or Mainland China from 18 January 2020 until 4 February 2020, when most of the border crossings were closed [9]. After 28 January, most the travellers from Mainland require 14 days self-quarantine. Quarantine and border control were the major control measures performed during this period in Hong Kong.

Assessing the effect of quarantine measures during this period in Hong Kong may provide an opportunity to understand how an outbreak can be suppressed (indicated by a reproductive number below one), by quarantine measures without extremely intensive interventions. However, challenge exists in assessing the effect on transmissibility in Hong Kong using classical transmission models, such as susceptible-infectious-recovered (SIR) or susceptible-exposed-infectious-recovered (SEIR) models, because many confirmed cases were identified as imported cases. To obtain the reproductive number, an indication of the transmissibility of the virus, the number of secondary infections of a given infected individual should be estimated. However, with many imported cases, the observed changes in numbers of cumulative cases are not totally due to those secondary infections happened locally. Because current models cannot distinguish the imported and local infections, overestimation of the reproductive number can be easily happened.

In order to estimate the reproductive number and other epidemiological parameters of COVID-19 in Hong Kong, here we developed a susceptible-exposed-infected-quarantined-recovered (SEIQR) meta-population model embedded with passenger data from Mainland China, that can stratify imported and local cases. We used Hong Kong as an example to demonstrate that the model can successfully recapture the transmission dynamics of both imported and local infections and estimate the reproductive number. Furthermore, the minimum timing and intensity of quarantine to suppress the outbreak were estimated.

## Materials & Methods

### Data collection

We collected the date of symptom onset time for each daily newly infected case of COVID-19 from 18 January to 29 February 2020 in Hong Kong from the Centre for Health Protection, Government of the Hong Kong Special Administrative [10]. Number of daily newly infected COVID-19 cases in Wuhan City and Mainland China outside Wuhan from 16 January to 29 February 2020 were collected from the National Health Commission of China [11]. Daily passenger data from Mainland China during the corresponding period were obtained from the Hong Kong Immigration Department [12].

### SEIQR Meta-population model

The meta-population model was fitted first to the data from Wuhan and Mainland China (outside Wuhan). Using the reconstructed transmission dynamics from source regions, the model was next fitted to the confirmed cases with symptom onset in Hong Kong with transportation data. Assuming the newly emergence of COVID-19 causes an outbreak at location *i*, during the emergence, the changes of the numbers of infectious cases *I*_*j*_ at a different location *j* can be determined using a meta-population framework with a mobility matrix (contact mixing at the population level) such that *I*_*j*_ = *I*_*j_imp*_ + *I*_*j_loc*_, where the subscripts *imp* and *loc* represent imported and local cases and the number of *I*_*j_imp*_ is dependent on the mobility matrix. We developed an SEIQR model to include dynamics of both imported and local cases at a target location (Hong Kong), and linked this model to the meta-population framework:

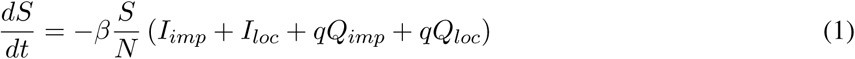

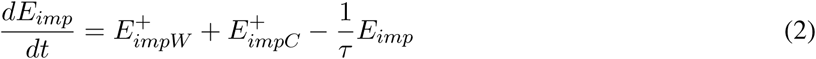

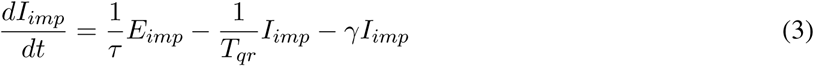

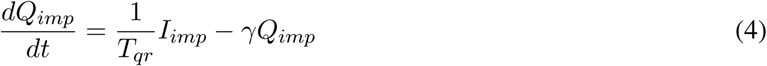

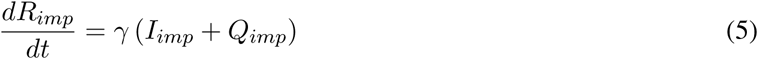

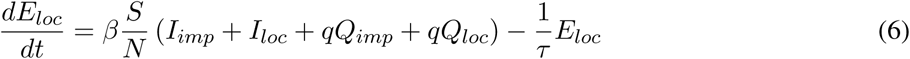

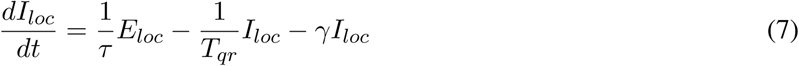

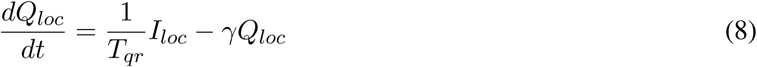

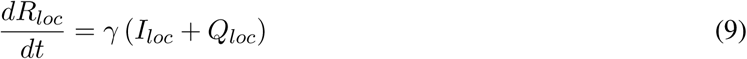

where *β* is the transmission rate, *τ* is the latent periods, *γ* is the recovery rate, *Tqr* is the time to quarantine after being infectious and *q* is the recontact ratio of quarantined to unquarantined individuals. *I*_*imp*_, *I*_*loc*_, *Q*_*imp*_, *Q*_*loc*_ are the infectious imported cases, infectious local cases, infectious imported cases that are under quarantined and infectious local cases that are under quarantined. Please see Table 1 and Table 2 for detailed definitions of each variables and parameters. Our SEIQR meta-population model stratifies the imported and local cases and is embedded with border control and quarantine measures given daily passenger data from Wuhan and Mainland China. 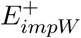 and 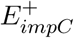 he daily newly number of imported cases from Wuhan (denoted as *W*) and Mainland China outside Wuhan (denoted as *C*). These numbers are determined by the daily passenger numbers and incubation period:

**Table 1:** Descriptions of variables in the meta-population model.

| Variables | Descriptions |
| --- | --- |
| $N$ | Population size in Hong Kong |
| $S$ | Susceptible cases |
| $E_{imp}$ | Exposed imported cases |
| $I_{imp}$ | Infectious imported cases |
| $Q_{imp}$ | Quarantined infectious imported cases |
| $R_{imp}$ | Recovered imported cases |
| $E_{loc}$ | Exposed local cases |
| $I_{loc}$ | Infectious local cases |
| $Q_{loc}$ | Quarantined infectious local cases |
| $R_{loc}$ | Recovered local cases |
| $N_W$ | Population size in Wuhan |
| $S_W$ | Susceptible cases in Wuhan |
| $I_W$ | Infected cases in Wuhan |
| $N_C$ | Population size in Mainland China (non-Wuhan) |
| $S_C$ | Susceptible cases in Mainland China (non-Wuhan) |
| $I_C$ | Infected cases in Mainland China (non-Wuhan) |

**Table 2:** Values of reproductive numbers and other epidemiological parameters of COVID-19 in Hong Kong. Mean values with 95% credible intervals are produced. The basic reproductive number is defined as the expected number of secondary infections without quarantine. The effective reproductive number is derived from the posterior variables when quarantine measure is considered. The incubation time *inc* refers to the sum of latent period *τ* and pre-symptomatic transmission period *σ*.

| Parameters | Values | Descriptions |
| --- | --- | --- |
| $R_e$ | 0.76 (0.66 - 0.86) | Effective reproductive number |
| $R_0$ | 2.32 (1.19 - 4.42) | Basic reproductive number (without quarantine measure) |
| *inc* | 5.43 (1.30-9.47) | Incubation time (unit: day) |
| $\tau$ | 1.94 (0.34-4.17) | Latent period (unit: day) |
| $\sigma$ | 3.49 (0.48-5.80) | Pre-symptomatic transmission period (unit: day) |
| $T_c$ | 7.80 (6.81-8.79) | Generation time (unit: day) |
| $T_{qr}$ | 2.92 (0.65-7.81) | Time to quarantine after being infectious (unit: day) |
| $q$ | 10.26% (5.32-17.8) | Recontact ratio of quarantined to unquarantined |
| $R_{pt}$ | 22% (9-41) | Reporting ratio in Mainland China |

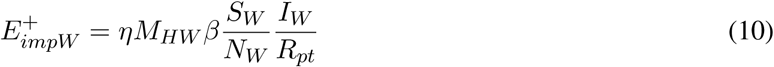

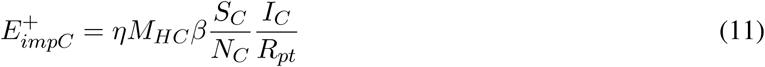

where *M*_*ji*_ is the mobility rate from *i* to *j*, subscripts *H, W, C* indicates Hong Kong, Wuhan, Mainland China (outside Wuhan), respectively. We used daily passenger data from the source regions divided by the population size in the source regions to refer to *M*_*ji*_ (Table S1). Among all the passengers from Mainland China, the proportion of them coming from Wuhan to Hong Kong during the study period can be calculated using the International Air Transport Association (IATA) database [13]. We estimated 2.92% of the total passenger from Mainland China to Hong Kong was from Wuhan. We assumed only the patients without symptom onset can pass the border to Hong Kong. Thus the terms 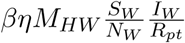 and 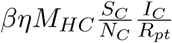 represent the daily newly imported cases from Wuhan and Mainland China (outside Wuhan) where *η* is a function to calculate the number of infected cases before symptom onset [14] and *R*_*pt*_ is the reporting ratio.

### Transmission dynamics in source regions

To obtain the number of imported cases, the model has to generate the transmission dynamics (*I*_*W*_ and *I*_*C*_) at source regions and estimated the imported cases using transportation data. We used a simple SIR model to construct newly infected numbers that reproduced the same numbers of cumulative confirmed cases in Wuhan and Mainland China (outside Wuhan) after the outbreak. Please see supplementary methods for the detailed descriptions.

### Parameter estimation

Prior to parameter estimation, the transmission dynamics in Wuhan and non-Wuhan Mainland China were reconstructed using a modified SIR model. The resulting source dynamics were used as an initial condition to seed imported cases for the target region (Figure S1 and Figure S2). Second, the posterior distributions of the parameters of transmission dynamics in Hong Kong were obtained using Markov Chain Monte Carlo (MCMC) to fit the detected infected cases from the model to the reported cases with symptom onset. The posterior distributions were obtained from Metropolis-Hastings in Markov Chain Monte Carlo (MCMC) with 1.2 × 10^6^ steps (Figure S3 and Figure S4) to guarantee an effective sample size (ESS) of greater than 200 for all parameters. The Gelman-Rubin convergence diagnostic was used and all the scores were less than 1.056 and near one, which confirmed the convergence.

Prior distributions for all the parameters were set to uniform distributions except the generation time *Tc* and the recontact ratio *q*. The prior of the generation time was assumed to be normally distributed with mean set to be 7.95 days, the average from two previous studies [15, 16]. The standard deviation was 0.25. The mean recontact ratio of being quarantined and not quarantined in a Gaussian prior was set to be 12% with a standard deviation of 0.05. A recent study has estimated each individual can contact 12.5 persons on average during a day [17]. Assuming many self-quarantine individuals can likely contact to their household members, with a possible expected number 1.5 persons, the mean recontact ratio of quarantined individuals to unquarantined in the prior distribution can thus be determined as 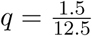.

### Likelihood of symptom onset

The likelihood of observing onset dates of both confirmed imported and confirmed local cases in Hong Kong were calculated based on Poisson distribution during MCMC. Newly detected infected cases are defined as the newly infected cases with symptom onset that eventually became quarantined. We assumed all the confirmed cases were quarantined. The daily newly detected imported cases with symptom onset was used as the expected value of the Poisson distribution and can be derived as Δ*I*_*imp*_(*σ*)*D*, where 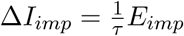, and *D* is the detection ratio, which was defined as the proportion of the number of quarantined imported cases to the total number of infectious and quarantined cases. *σ* represents the pre-symptomatic transmission period, which produced a delay of onset after an individual has been infected. The same approach can be used to determine the expected value of the Poisson distribution for the detected local cases.

### Effective reproductive number calculation

The effective reproductive number *R*_*e*_, was calculated using the next-generation matrix approach after obtaining the posterior distributions of model parameters [18]. Please see supplementary methods for the detailed descriptions.

## Results

### Dynamics of imported and local cases

Our meta-population model reproduced the COVID-19 transmission dynamics of both imported and local infections in Hong Kong. The number of cumulative imported cases was increasing rapidly in Hong Kong after the first imported case was detected, with onset day on 18 January 2020 until the end of Chinese New Year in early February (Figure 1A). The risk of community spread was highlighted as the number of local cases crossed above the imported cases. In order to understand the transmission dynamics of COVID-19 in Hong Kong, we developed an SEIQR meta-population model that stratifies imported and local cases (Figure 2). The model recaptured the cumulative numbers with a crossover between the local and imported cases (Figure 1B) and transient dynamics (Figure 3). The predicted number of imported cases reached to a peak on 26 January (Figure 3A). These imported cases immediately caused a wave of local infections. The number of daily newly detected local cases reached to a peak around 2 February and decreased afterward (Figure 3B).

**Figure 1.**
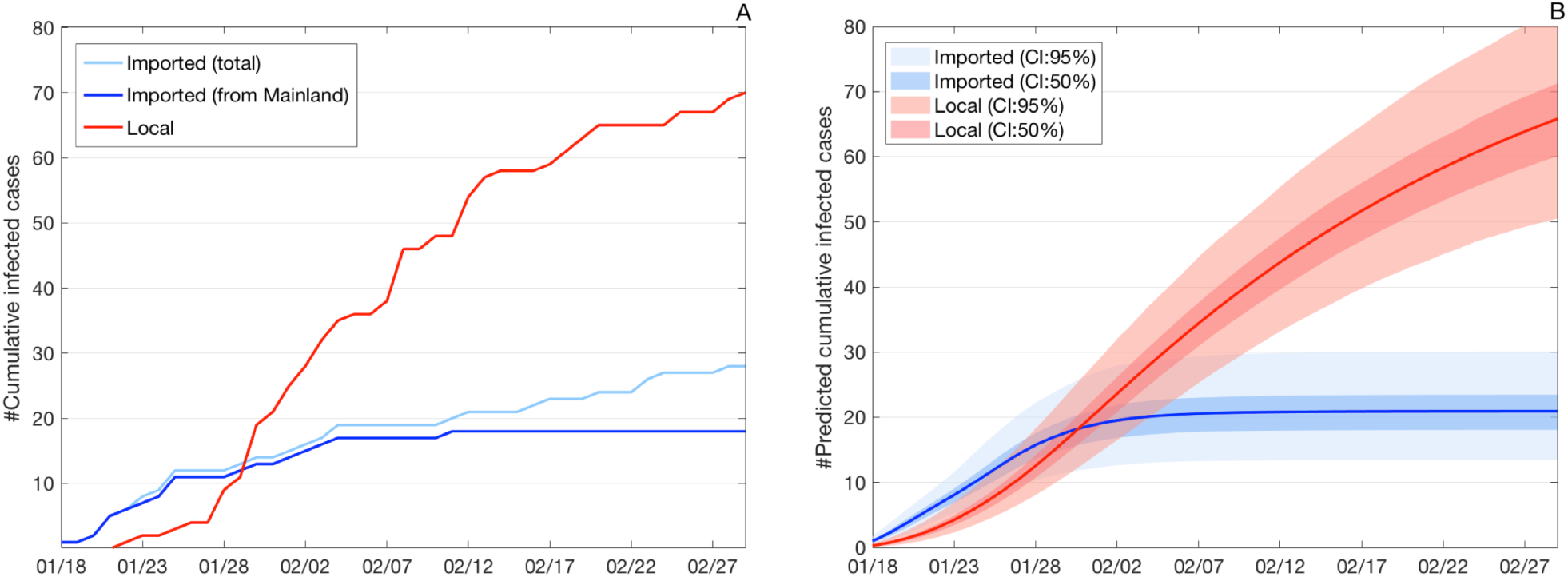
Number of cumulative confirmed COVID-19 cases by symptom onset date. (A) Observed cumulative confirmed cases. Light blue denotes the total imported cases. Dark blue denotes the imported cases only from China, excluding few other cases mainly from diamond princess cruise. Red denotes the local cases. (B) Predicted cumulative detected cases. Blue denotes the predicted number of cumulative detected imported cases from China. Red denotes the predicted number of cumulative detected local cases. Solid lines indicate the mean values.

**Figure 2.**
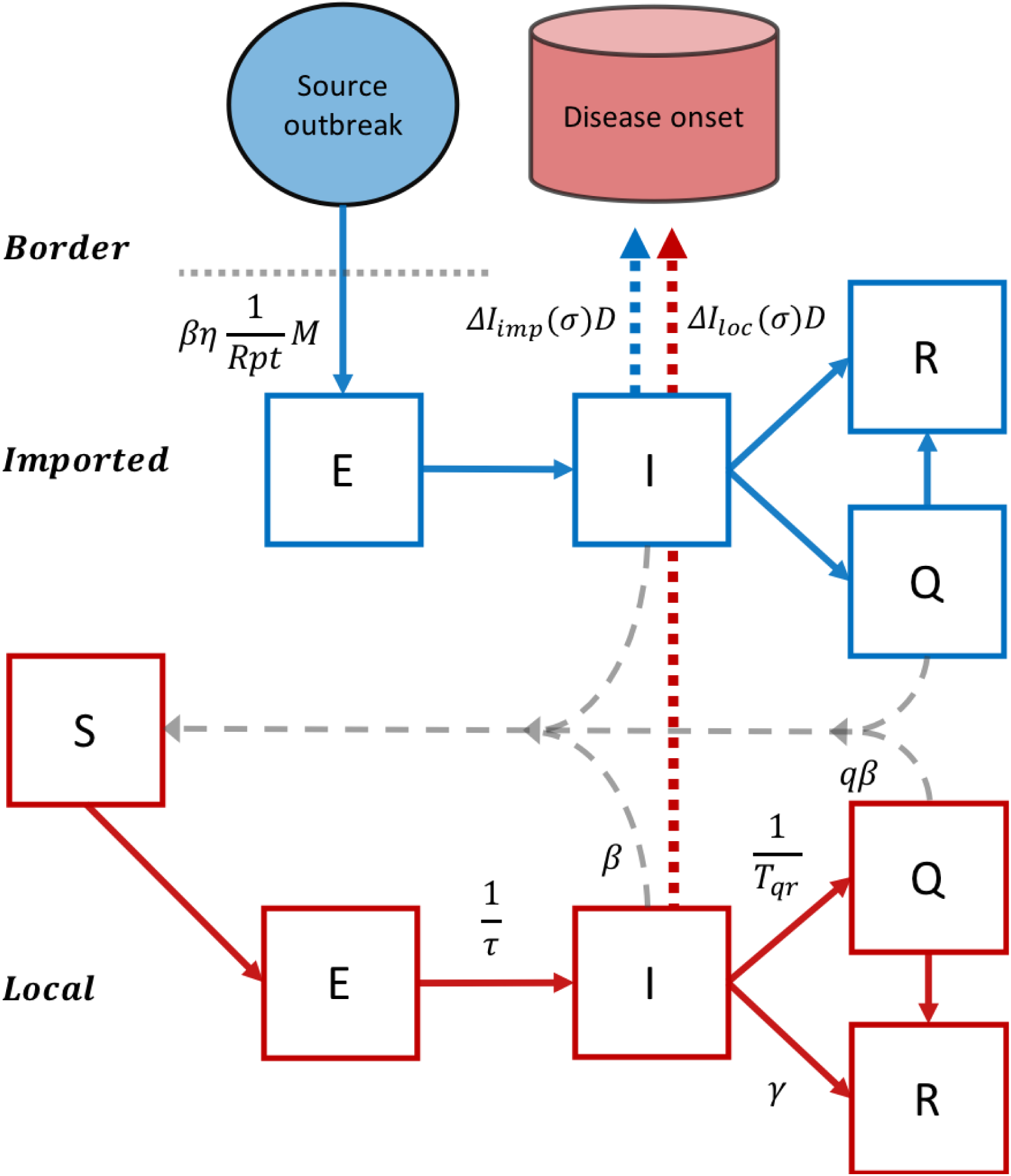
Schema of susceptible-exposed-infected-quarantined-recovered meta population model. Imported cases arrive as exposed (E) status before disease onset in order to pass through the border. Imported cases then become infectious (I), quarantined (Q) or recovered (R) statuses. 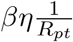 *M* is the rate to produce imported cases, where *η* is a function to determine the probability of an ill passenger can pass the border (see Methods for the details). *R*_*pt*_ is the reporting ratio in Mainland China and *M* is the mobility rate. Both imported and local cases are able to infect susceptible individuals (S) and cause local transmission while quarantined cases have a lower transmission rate depending on the recontact ratio (*q*), indicating the ratio of the contact rates of quanratined to unquarantined individuals. Epidemiological parameters *β* is the transmission rate, *τ* is the latent periods, *γ* is the recovery rate, *T*_*qr*_ is the time to quarantine after being infectious. Δ*I*_*imp*_(*σ*)*D* represents the newly detected imported cases, where *D* is the detected ratio defined as the ratio of the number of quarantined imported cases to the total number of infectious and quarantined imported cases, and *σ* is the pre-symptomatic transmission period, indicating the delayed time of symptom onset after being infectious. Similarly, Δ*I*_*loc*_(*σ*)*D* represents the detected local cases.

**Figure 3.**
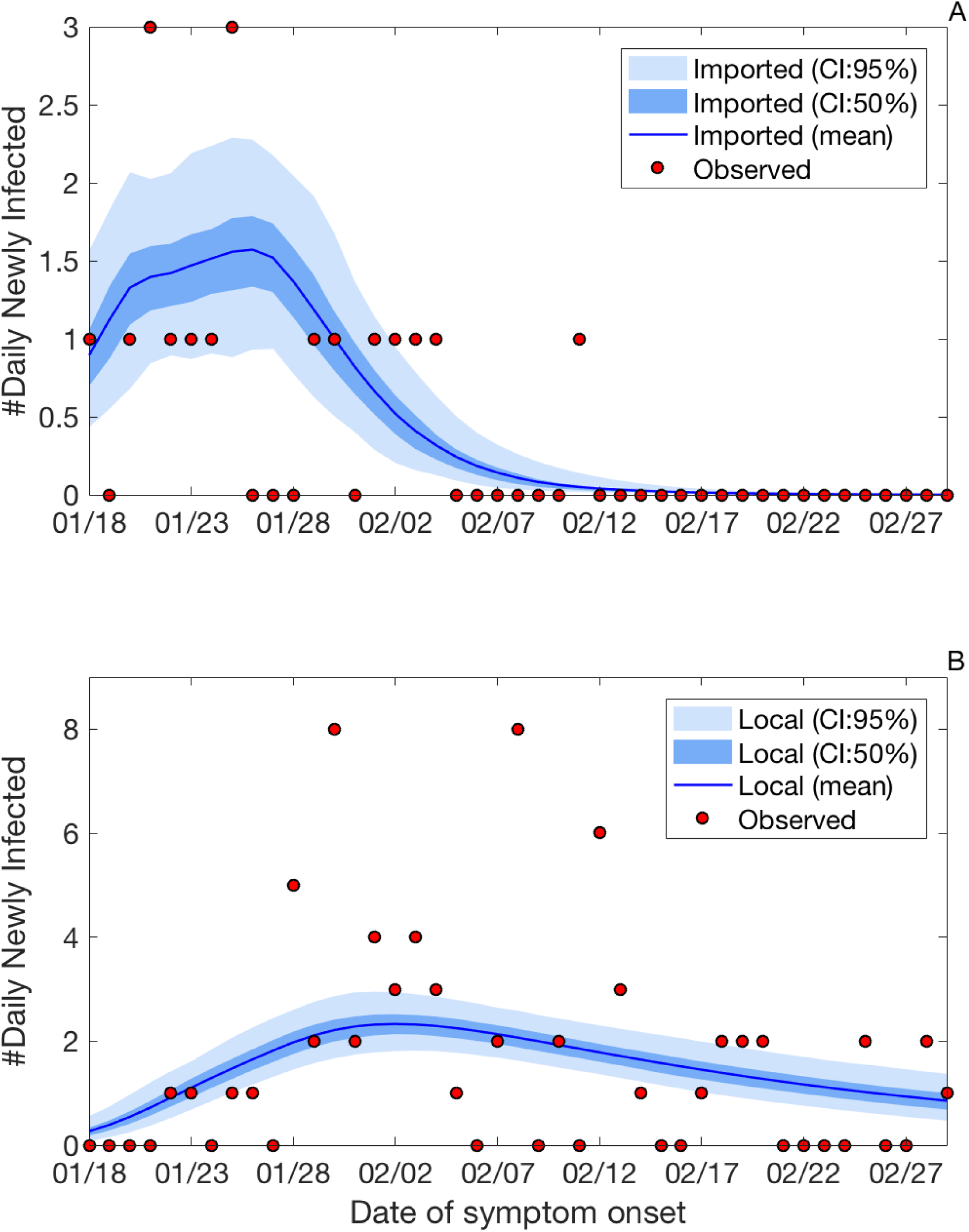
Observed and predicted numbers of detected imported and local cases. (A) Number of observed imported and predicted number of detected imported cases. Mean, 50% and 95% prediction intervals are shown. The predicted detected cases are the daily newly cases with symptom onset that eventually become quarantined and detected. (B) Number of observed local and predicted number of detected local cases. Mean, 50% and 95% prediction intervals are shown. The definition of the detected local cases is same as (A) but for local infections.

### Epidemiological parameters

Epidemiological parameters of COVID-19 were obtained for Hong Kong from 18 January to 29 February. The effective reproductive number *R*_*e*_ was 0.76 (0.66 - 0.86) (Table 2). *R*_0_ was 2.32 (1.19 - 4.42), referring to the reproductive number when the quarantine measure was not included. Latent period was 1.94 days (0.34 - 4.17). The pre-symptomatic transmission period before disease onset was 3.49 days (0.48 - 5.80). The incubation time was calculated as 5.43 days (1.30 - 9.47), consistent with recent findings, where a mean or median incubation period of approximately 5 days was reported [16,19,20]. We assumed all the infected persons can pass the border before disease onset, which was defined as the incubation time. The estimated time to quarantine after being infectious *T*_*qr*_ in Hong Kong is 2.92 days (0.65 - 7.81) days, which is nearly two days less than the incubation period. The recontact ratio of quarantined to unquarantined individuals *q* was 10.26% (5.32 − 17.8). The model estimated only 22% (9 − 41) of total infected cases were documented in Mainland China.

### Effect of quarantine on Reproductive Number

The timing of quarantine was an important measurement to determine the risk of community spread. The estimated *T*_*qr*_ in Hong Kong was about half day earlier than the symptom onset time and was able to reduce *R*_*e*_ to be less than one (Figure 4A). If *T*_*qr*_ was longer than one day after disease onset, *R*_*e*_ became greater than one, the community spread could happen. Furthermore, the recontact ratio of quarantined to unquarantined individuals affected the value of *R*_*e*_. If the *q* became about two and a half larger than the estimated value, *R*_*e*_ was greater than one and the community spread could also happen (Figure 4B). The epidemiological parameters and the effects of quarantine measures during an infection generation was illustrated (Figure 5). Sensitivity analysis has been performed to evaluate the impact of initial setting of generation time to the effective reproductive number. For sensitivity analysis, we have tested two alternative model settings, with the generation time fixed at 7.5 or 8.4 days. Both of the settings gave the mean *Re* of 0.76 or 0.77 as well.

**Figure 4.**
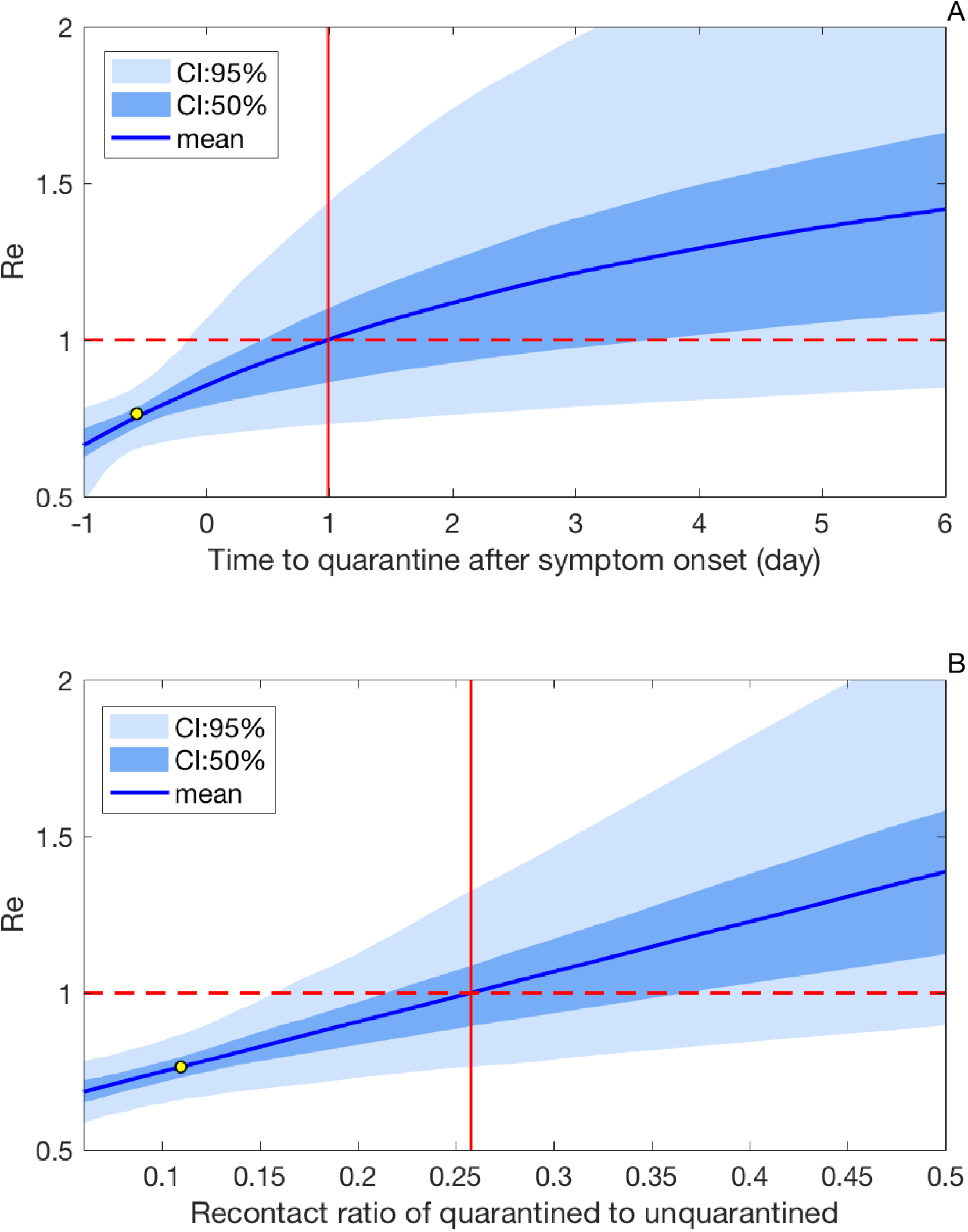
Effective reproductive numbers by different timings and intensities of quarantine measures. (A) The mean and credible intervals of effective reproductive number *R*_*e*_ by different days of time to quarantine *T*_*qr*_. Note the values were adjusted to time to quarantine after symptom onset. The vertical line corresponds to the average time from onset to isolation when *R*_*e*_ is one. Dashed line indicates the level of *R*_*e*_ as one. Yellow dot represents the estimated *T*_*qr*_ and the corresponded *R*_*e*_. (B) The mean and credible intervals of effective reproductive number *R*_*e*_ by recontact ratios *q*. Yellow dot represents the estimated *T*_*qr*_ and the corresponded *R*_*e*_. All the values were estimated from 2000 random samples from posterior distributions.

**Figure 5.**
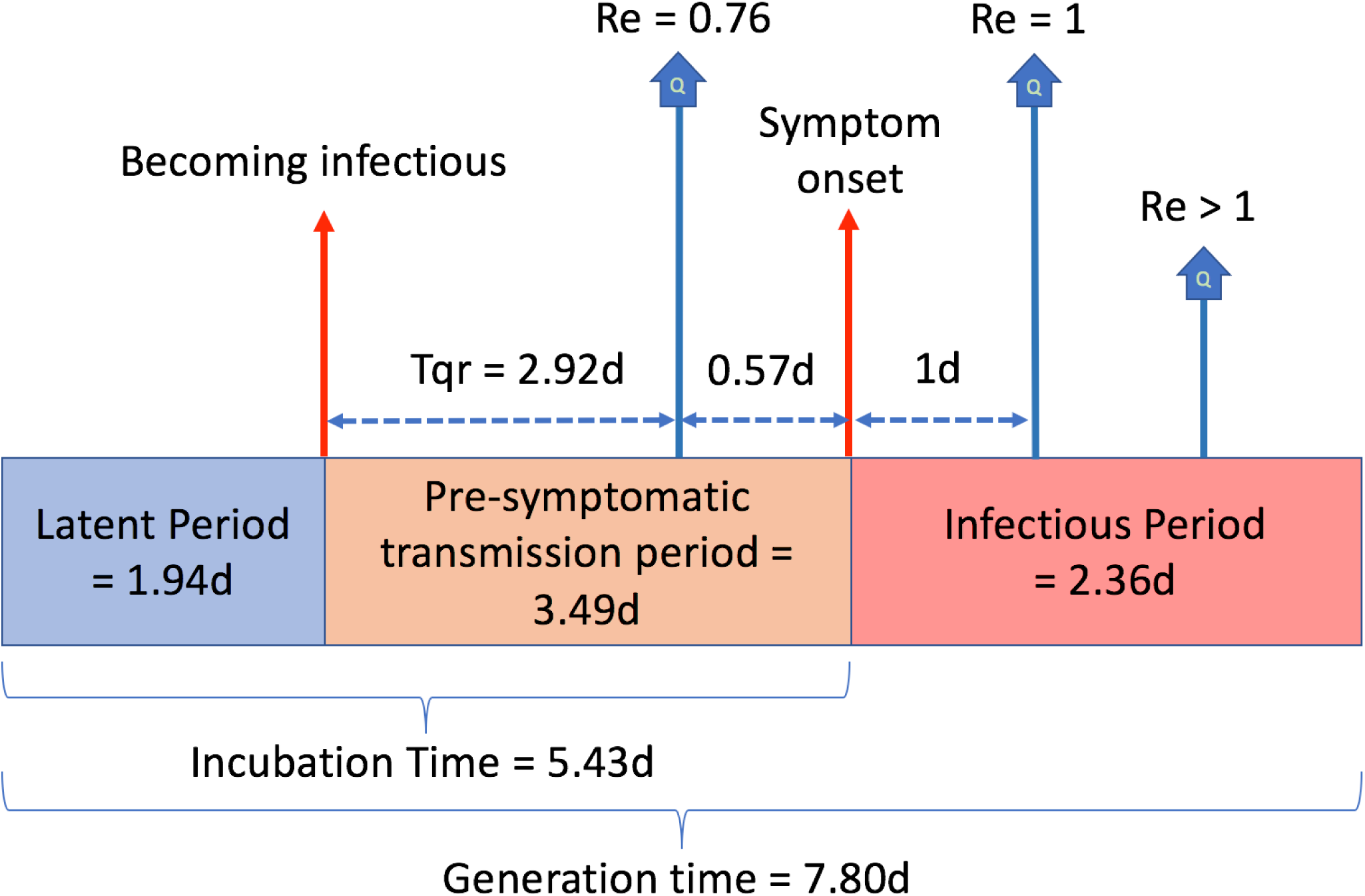
Epidemiological parameters during an infection generation. The effects of different times to quarantine on the effective reproductive number are illustrated.

### Effects of quarantine on Detection Ratio

The results showed that the ratio can be low during the initial period but soon reached saturated values after 3 weeks both for imported and local cases (Figure 6AB). Generally the detection ratios of the imported cases were higher than the local cases because the number of the imported cases were low. For local cases, 71%(46 − 90) of which were detected, estimated by our model. One day delay of quarantine reduced about 10% of daily detection ratio to 60%(39 − 74) (Figure 6B). Only 31%(20 − 39) of them could be detected or quarantined if quarantine was delayed 6 days. The results showed that not only early quarantine can reduce reproductive number, but also has a benefit on increasing overall detection ratio.

**Figure 6.**
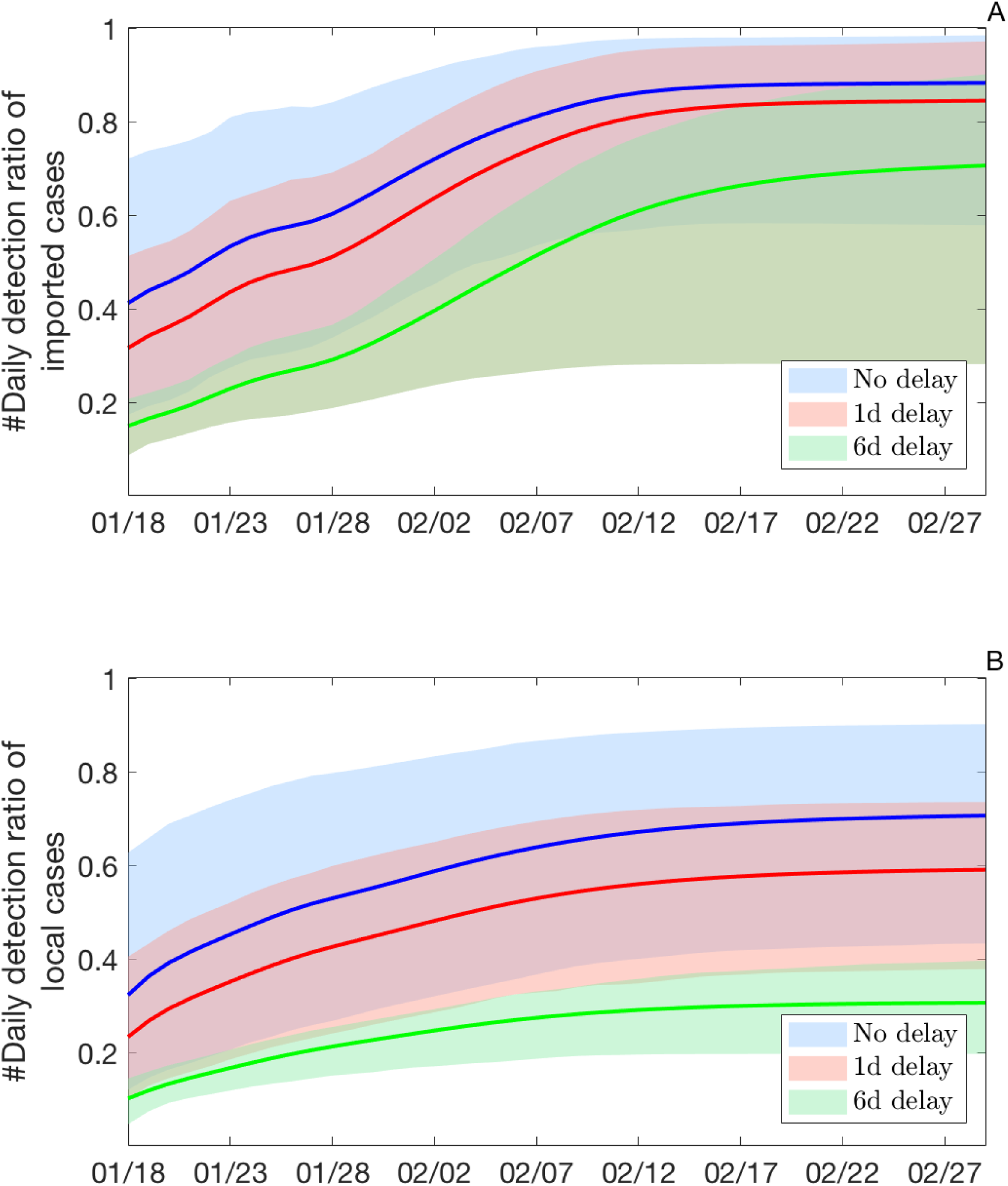
Detection ratios of imported and local cases by time. (A) Detection ratios of imported cases. The ratios denote the proportion of daily newly cases with symptom onset that eventually become quarantined among all infectious cases. Blue, the detection ratio estimated using the posterior distribution of the time to quarantine, denoted as no delay. Red, the detection ratio estimated using the posterior distribution of the time to quarantine with one day delay. Green, the detection ratio estimated using the posterior distribution of the time to quarantine with six days delay. Shared areas are the 95% intervals. (B) Detection ratios of local cases. The ratios denote the proportion of daily newly cases with symptom onset that eventually become quarantined among all infectious cases. Same definition of colors are used as (A) but for local infections.

## Discussion

By characterizing the transmission dynamics with quarantine and border control measures across the Chinese New Year festival in Hong Kong, we have identified the key aspects in containing the outbreak. This is the first study to demonstrate that early time targeted quarantine measures significantly suppress the COVID-19 outbreak and the proper timing of quarantine is feasible. Suppression of the outbreak during the study period is important because global expansion occurred starting from this critical moment when initial travellers who carried the diseases moved to different countries [21–24].

Until now, how to suppress the outbreak of COVID-19 has been studied only in regions with many infections, such as Wuhan or China [4, 21, 25, 26]. Although certain strict public health policies can suppress the outbreak, these approaches can cause profound social and economic impacts, which may not be feasible by every country. In contrast, our study illustrated that Hong Kong can be a good model to learn how to prevent the community spread through quarantine before many other intensive control policies (such as transportation restriction and closure of public facilities) are required.

How to impose a large scale quarantine properly to suppress the outbreak remain largely unknown [6]. Our results demonstrated that quarantine of suspected individuals before symptom onset, is critical to contain the COVID-19 outbreak. Given that the incubation period was about 5 days and the confirmation of COVID-19 infection can often be delayed, to guarantee an early quarantine of all suspected cases before symptom onset is a critical criterion to reduce the chance of community spread.

Using a model that can stratify both dynamics of imported and local infections, we demonstrated that many epidemiological parameters, including the reproductive number, latent period and incubation time, along with the reporting ratio in Mainland China can be estimated after fitting confirmed cases with symptom onset in Hong Kong with daily transportation data. Generation time we used are adopted from previous studies studies with the estimated mean of 7.8 days [15, 16]. The values, such as incubation time and reporting ratio are consistent to recent studies [7, 16, 19, 20].

One of the major challenges to limit the COVID-19 epidemic through quarantine is pre-symptomatic transmission. Currently, little is known about when does the transmission occur before the symptom. Our model has found, an individual after latent period, can be infectious without symptoms about 3 days, similar to 1-3 days found by many recent studies through contact tracing and enhanced investigation of clusters of confirmed cases [27–30]. Thus, even if a very intensive contact tracing is proposed, to identify all the contacts made by an infectious individual before symptom onset can be difficult. Thus, how to reduce social contacts and maintain social distancing along with quarantine measures become critical to prevent the spread. To evaluate the benefits of different approaches to reduce unnecessary contacts, such as working from home, wearing masks, etc, becomes an important task [31, 32].

The study highlights the importance of having a time targeted quarantine measure along with travel bans. Investigating the timing and the quality of quarantine in different outbreak regions is a critical factor for prevention and control of COVID-19 outbreak.

## Data Availability

Data are publicly available.

## Acknowledgments

We thank Prof. Steven Riley from MRC Centre for Global Infectious Disease Analysis at Imperial College London to give valuable comments. We thank Prof. Mengsu (Michael) Yang, Dr. Xin Wang, and Dr. Kai Liu from City University of Hong Kong, Prof. Kin On Kwok from Chinese University of Hong Kong, Prof. Dong-Ping Wang from Stony Brook University, Dr. Lindsey Wu from London School of Hygiene and Tropical Medicine, and all the anonymous readers who have provided invaluable comments. The authors also acknowledge the support from the grants funded by City University of Hong Kong [#7200573 and #9610416].

## Author Contributions

H-YY designed the study. AM and GH participated in the data collection. H-YY and AM analysed and interpreted the data. H-YY wrote the paper. Everyone reviewed, revised and edited the manuscript.

## Supplementary Methods

### Dynamics in source regions

To obtain the number of imported cases, the model has to generate transmission dynamics in source regions is to seed the target region (Hong Kong). We modified an SIR to construct newly infected numbers that were close to the observed confirmed numbers in Wuhan and Mainland China (outside Wuhan).

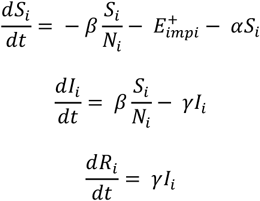

Where *α* is a parameter to represent the effect of reduction of social contacts after the Wuhan lockdown on 23 January 2020 and the closure of the border crossings on 4 February 2020 [1]. As a result, reduction of susceptible population happens by time. Assuming generation time is 8.4 days thus we have the recovery rate 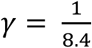. Using *R*_0_ = 2.92, we can obtain *β* = *R_0_ γ* = 0.3476 in Mainland China. We also assumed the recovery rate is same for *γ* both transmission dynamics in Mainland China and in Hong Kong.

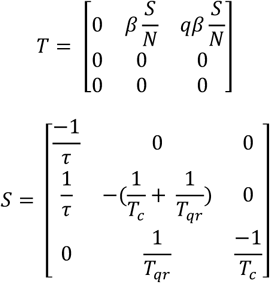

### Effective reproductive number calculation

The effective reproductive number *R*_*e*_, was calculated using the next-generation matrix approach after obtaining the posterior distributions of model parameters. Following the same notation as in the study by Diekmann et al[2]. We obtained the transmission matrix *T* and the transition *S*. Elements in *T* represents the average newly infected cases in exposed group (E) transmitted by a single infected individual in infectious or quarantined group (I), which can be calculated as 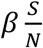 or 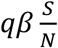. *R* can be calculated as the first eigenvector of −(*TS*^−1^) with the following formulas:

## Supplementary Figures and Tables

**Figure S1.**
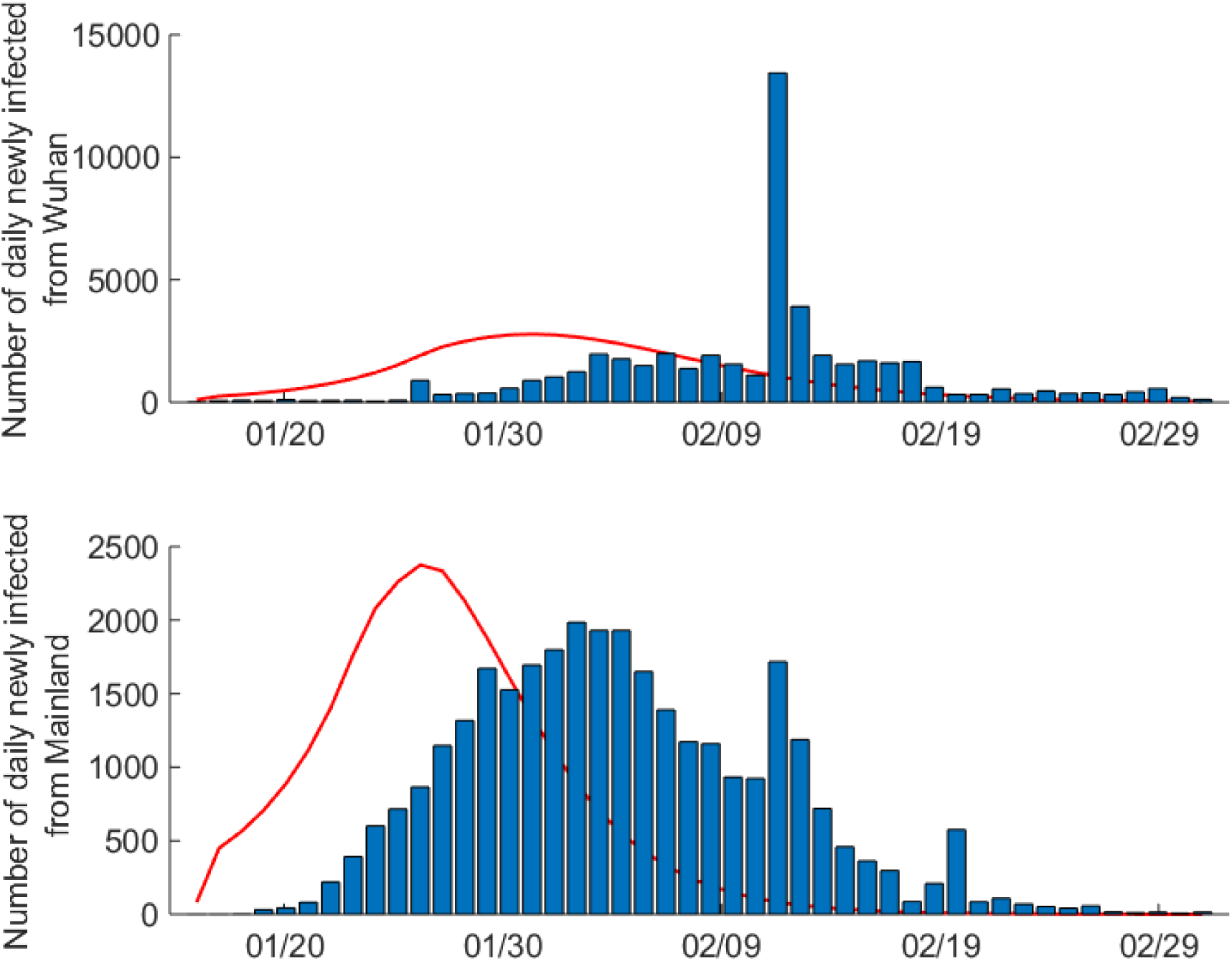
The number of daily newly infected COVID-19. cases by onset day in China. (A) Daily newly infected COVID-19 cases in Wuhan, China. Red line represents the reconstructed curve adjusted by 10 days reporting delay after fitting the actual data in Wuhan. The x-axis denotes the case reporting day after Jan 16, 2020. (B) Daily newly infected COVID-19 cases in Mainland China (outside Wuhan). Red line represents the reconstructed curve adjusted by 10 days reporting delay after fitting the actual data outside Wuhan.

**Figure S2.**
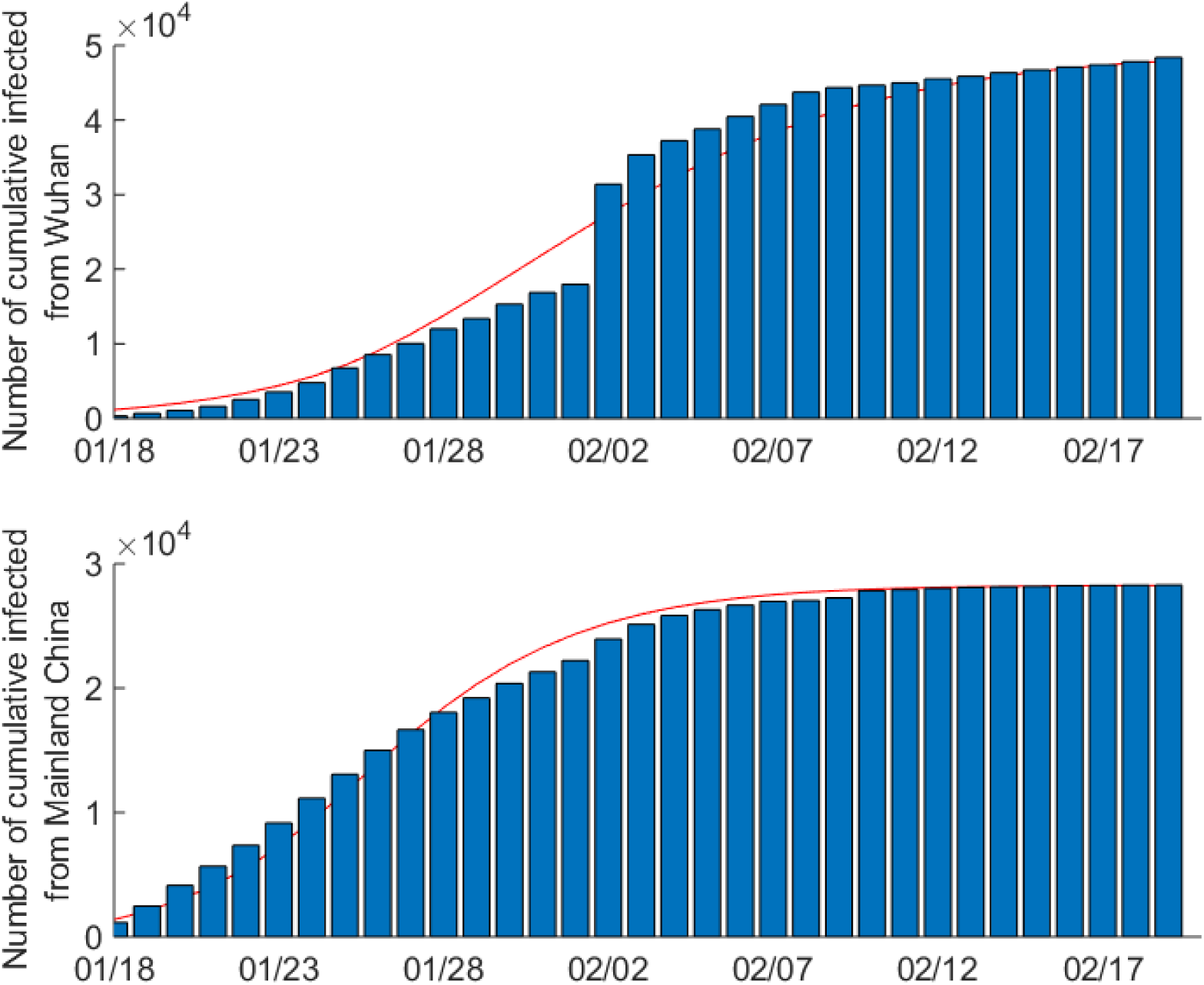
The number of cumulative confirmed COVID-19 cases by onset day in China. (A) Cumulative confirmed COVID-19 cases in Wuhan, China. Red line represents the reconstructed curve adjusted by 10 days reporting delay after fitting the actual data inside Wuhan. The x-axis denotes the number of days from Jan 18 to Feb 29, 2020. (B) Cumulative confirmed COVID-19 cases in Mainland China (outside Wuhan). Red line represents the reconstructed curve adjusted by 10 days reporting delay after fitting the actual data outside Wuhan.

**Figure S3.**
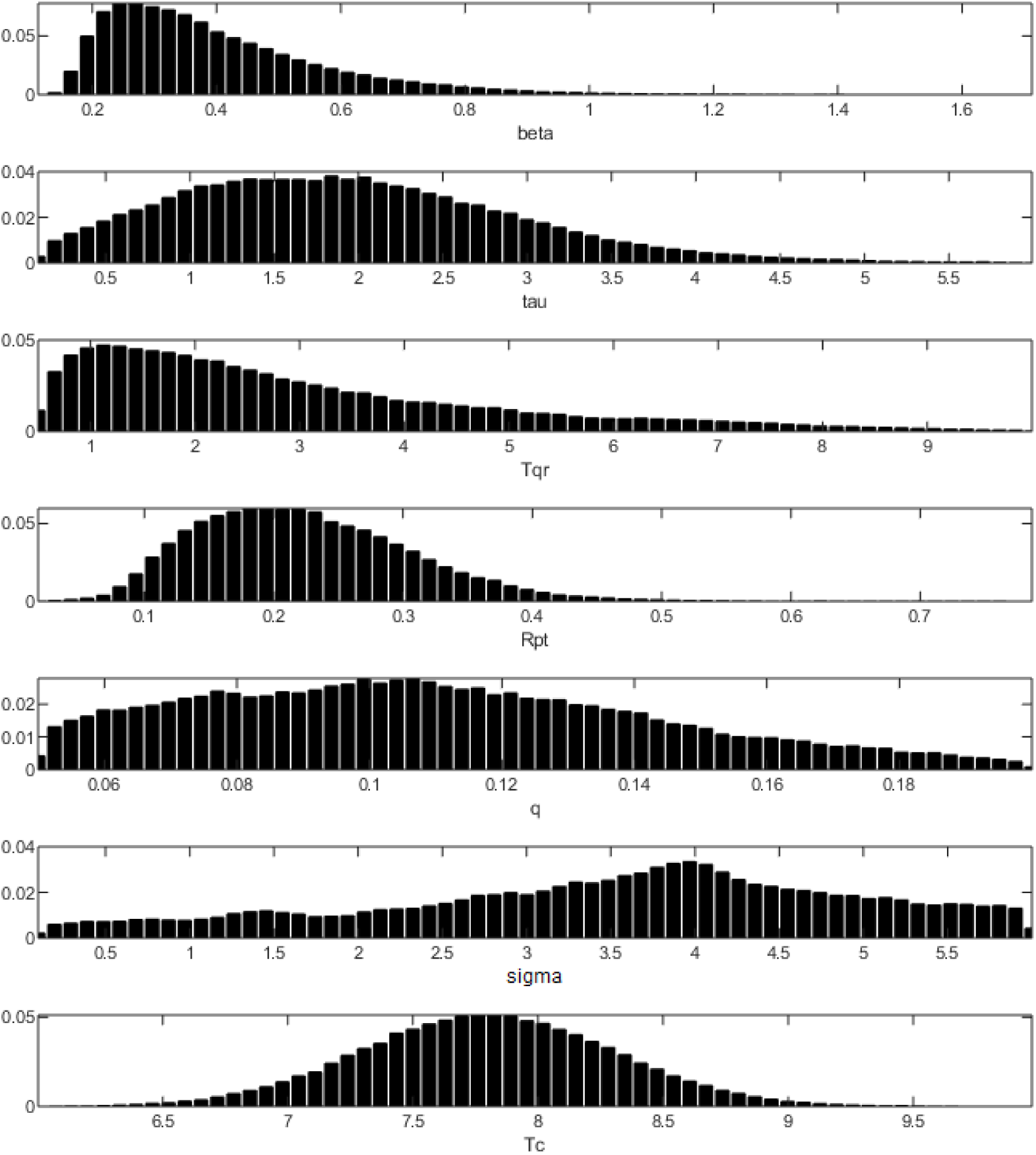
Posterior distributions of model parameters.

**Figure S4.**
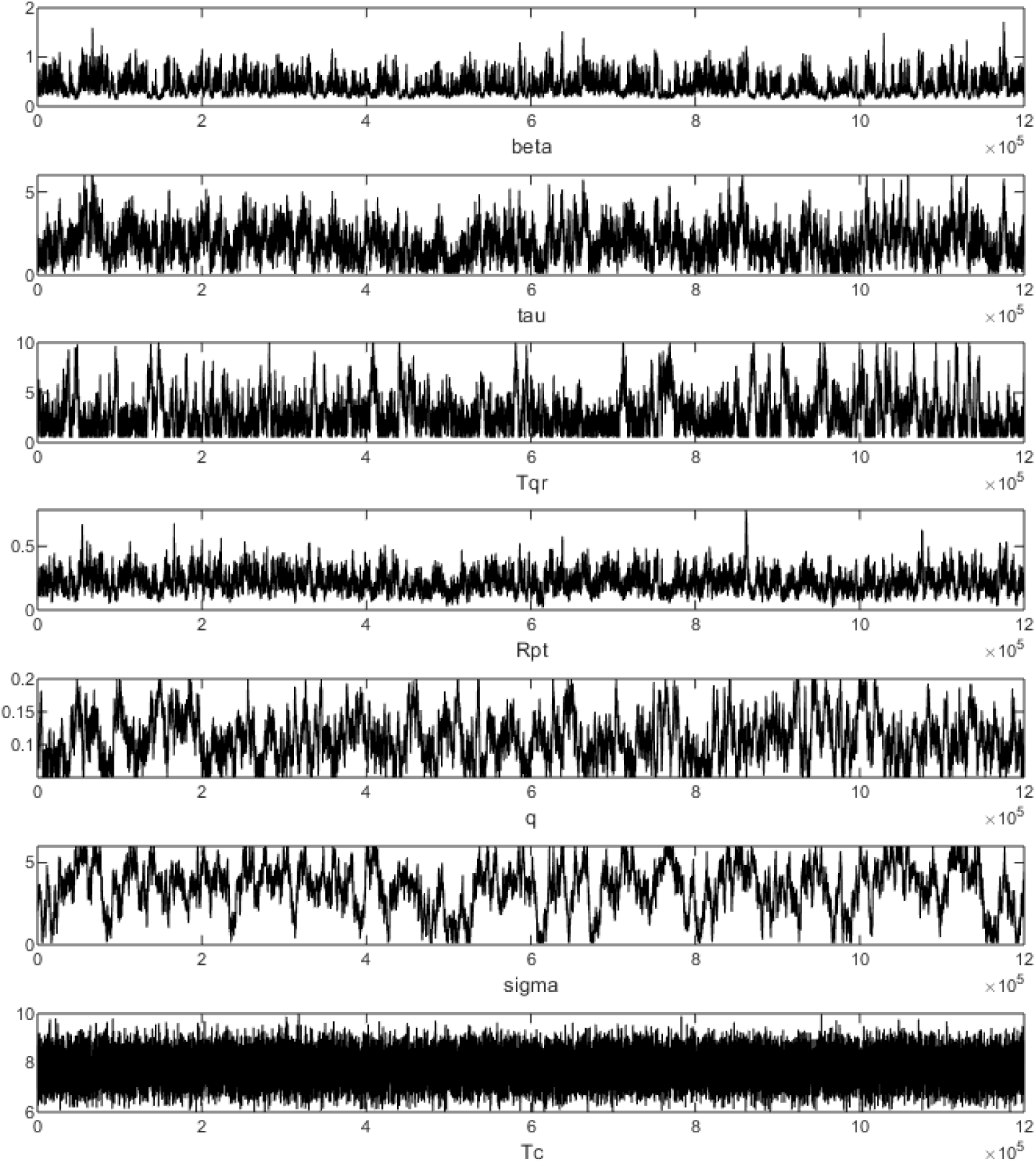
Trajectories of MCMC output.

**Table S1.** Daily number of passengers from outside Hong Kong

| **Time** | **Residents(HK)** | **Residents(Mainland)** | **Time** | **Residents(HK)** | **Residents(Mainland)** |
| --- | --- | --- | --- | --- | --- |
| 1/24 | 102,663 | 36,705 | 2/12 | 15,792 | 729 |
| 1/25 | 86,417 | 29,891 | 2/13 | 13,312 | 751 |
| 1/26 | 163,668 | 36,690 | 2/14 | 13,507 | 683 |
| 1/27 | 177,470 | 28,780 | 2/15 | 16,089 | 791 |
| 1/28 | 188,788 | 24,156 | 2/16 | 20,325 | 749 |
| 1/29 | 197,572 | 27,780 | 2/17 | 15,990 | 643 |
| 1/30 | 132,506 | 19,555 | 2/18 | 14,383 | 579 |
| 1/31 | 116,544 | 16,058 | 2/19 | 12,801 | 567 |
| 2/1 | 115,122 | 13,382 | 2/20 | 13,677 | 648 |
| 2/2 | 122,399 | 11,715 | 2/21 | 14,557 | 682 |
| 2/3 | 111,033 | 13,461 | 2/22 | 15,440 | 651 |
| 2/4 | 54,816 | 9,511 | 2/23 | 20,241 | 673 |
| 2/5 | 44,566 | 8,760 | 2/24 | 16,972 | 588 |
| 2/6 | 65,122 | 11,009 | 2/25 | 15,207 | 558 |
| 2/7 | 76,899 | 12,746 | 2/26 | 14,317 | 587 |
| 2/8 | 18,823 | 995 | 2/27 | 13,846 | 702 |
| 2/9 | 24,939 | 956 | 2/28 | 15,248 | 558 |
| 2/10 | 18,408 | 832 | 2/29 | 17,881 | 1,084 |
| 2/11 | 15,953 | 738 |  |  |  |

